# Integrating Group and Individual Fairness Auditing in Clinical AI: A Post-Hoc, Model-Agnostic Approach

**DOI:** 10.1101/2025.09.03.25334999

**Authors:** Javen Xu, Yeon-Mi Hwang, Samhita Kondareddy, Inés Dormoy, Serena Liang Jing, Malvika Pillai, Catherine Curtin, Tina Hernandez-Boussard

## Abstract

**Objectives:** To develop and demonstrate a practical post-hoc, model-agnostic fairness auditing tool that integrates group-level and individual-level fairness assessment for clinical prediction models.

**Materials and Methods:** We developed EquiLense, a fairness auditing tool that operationalizes three components: group fairness evaluation using established and novel group-level disparity metrics, individual fairness assessment, and the Mean Predicted Probability Difference (MPPD), a novel metric quantifying prediction inconsistencies between clinically similar patients across demographic groups. We applied EquiLense to EHR data from 59,047 surgical patients across post-surgical delirium and 30-day readmission prediction models, and demonstrated MPPD on two external benchmarks with documented bias: the COMPAS recidivism dataset and the UCI Adult Income dataset.

**Results:** Applied to delirium prediction (AUROC 0.78), MPPD identified prediction inconsistencies between clinically similar patients that varied systematically across demographic groups. Among patients clinically similar to White patients, Asian patients exhibited the largest disparity (MPPD difference = 0.045). In external benchmarks, MPPD assigned the highest disparity scores to African American defendants in COMPAS and to female workers in UCI Adult Income, consistent with documented bias in both datasets.

**Discussion:** Current fairness evaluation approaches address group-level disparities or individual-level consistency in isolation, creating blind spots in algorithmic fairness assessment. EquiLense bridges this gap by integrating established fairness metrics with MPPD in a single post-hoc tool, enabling comprehensive and interpretable fairness auditing without model retraining.

**Conclusion:** EquiLense provides a practical approach to fairness auditing in clinical AI by combining group and individual fairness perspective through MPPD, a novel metric demonstrated on both clinical and benchmark datasets.

**Lay Summary:** Clinical prediction models are increasingly used across clinical care, research, and decision-making, but they can produce unfair outcomes for patients. Fairness can be evaluated at the group level, by comparing outcomes across demographic categories, or at the individual level, by assessing whether clinically similar patients receive consistent predictions. Most existing tools address one perspective but not both together. We developed EquiLense, a post-hoc fairness auditing tool that bridges these two perspectives. EquiLense operationalizes three components: (1) group fairness evaluation using using multiple group-level disparity metrics; (2) individual fairness assessment measuring prediction consistency among clinically similar patients; and (3) the Mean Predicted Probability Difference (MPPD), a novel metric that quantifies how differently a model treats two patients who are clinically similar but belong to different demographic groups. We applied EquiLense to surgical outcome prediction models using data from over 59,000 patients, and validated MPPD on two external datasets with well-documented bias (COMPAS recidivism and UCI Adult Income). MPPD detected known disparities in both benchmarks. In our clinical application, we found that models can achieve strong overall performance while still producing systematically different predictions for clinically similar patients from different sensitive attributes. EquiLense requires no model retraining, applies to any clinical prediction model, and produces interpretable outputs grounded in clinically familiar features, making it directly applicable in real-world healthcare settings.

## 1. BACKGROUND AND SIGNIFICANCE

As artificial intelligence (AI) becomes increasingly integrated in clinical decision-making, ensuring algorithmic fairness is both an ethical priority and a practical challenge.[1–5] Current fairness evaluations are fragmented, focusing either on group-level disparities (e.g. across demographic categories) or individual level inconsistencies.[2, 6] This creates blind spots. Group metrics can obscure important patient-level variations, while individual metrics may fail to reveal systemic biases affecting entire demographic groups.[7]

Group-level metrics are valuable for identifying systemic disparities that disadvantage marginalized populations. However, they cannot reveal whether clinically similar individuals across demographic groups receive consistent predictions.[2, 8, 9]

Individual fairness frameworks address this gap by ensuring prediction consistency among comparable patients. Dwork et al.[10] define individual fairness as a constraint that any two similar individuals, as defined by a task-specific metric over observable features, should receive similar predictions. This is distinct from group fairness. A model can show no group-level disparity while still treating clinically similar individuals inconsistently across demographic groups. Although it is intuitively appealing, individual fairness has seen limited adoption in healthcare, where research remains in its early stages.[11] A key challenge is that defining a clinically meaningful similarity metric is inherently subjective, as different clinical contexts and tasks may require different definitions of similarity. Additionally, data-driven similarity metrics risk encoding existing biases, which may perpetuate rather than reduce health inequities.[11, 12] Despite growing recognition of its importance, practical tools for systematically quantifying individual-level fairness in healthcare remain limited.[5] Counterfactual fairness (CF) operationalizes a related but distinct principle through a causal lens, asking whether a prediction would change if the sensitive attribute were different, but requires causal graph specification or synthetic counterpart generation that is difficult to justify in EHR settings.[11, 13–15]

Recognizing that group and individual fairness each capture only partial perspectives, several approaches have sought to integrate both. Speicher et al.[16] introduced a generalized entropy-based metric that decomposes total unfairness into between-group and within-group components, offering a unified measure of disparity. Xu and Strohmer [6] presented a theoretical framework that characterizes the conditions under which group and individual fairness are compatible, but without operational implementation. FairGI jointly optimizes both using adversarial learning for graph neural networks.[17] In our prior work, FairEHR-CLP[18] combines CF-inspired individual fairness through synthetic counterpart generation with group fairness evaluation, jointly optimized during model training using contrastive learning. However, some of these approaches lack operational implementation, while others require model retraining or are tied to specific architectures, limiting their applicability for auditing already-deployed clinical models, a gap that EquiLense is designed to fill.

We introduce EquiLense, a post-hoc, model-agnostic fairness auditing tool that integrates group fairness evaluation, individual fairness assessment, and MPPD, a novel metric designed to bridge group-level disparity detection and individual-level prediction consistency, into a single accessible tool. Existing metrics each capture important but partial perspectives on fairness. EquiLense brings them together, with MPPD as its methodological contribution.

MPPD quantifies prediction inconsistencies between real, observed patients who are clinically similar but belong to different demographic groups. It operationalizes consistency-based individual fairness by grounding similarity in observable clinical features using cosine similarity, making its assumptions explicit and directly adjustable by practitioners.[10, 11] We demonstrate EquiLense using real-world surgical outcome prediction models and evaluate MPPD on two external benchmarks with known bias.

## 2. MATERIALS AND METHODS

### 2.1 Study design and population

This study was approved by the Institutional Review Board at Stanford University (Stanford, CA, USA; Protocol IRB-34551) with a waiver of informed consent. We applied Equilense to EHR data from an integrated healthcare system operating on a unified Epic platform. The cohort included 59,047 surgical patients treated between January 1, 2012 to December 1, 2022. The cohort included patients aged 50 years or older who had inpatient stays of 90 days or less and survived at least 30 days post-surgery. Our analysis incorporated all post-surgical clinical documentation, including progress and nursing notes. For patients with multiple surgeries, only the initial procedure was included. Cohort details are available in Table 1, Figure S1 and described in detail elsewhere.[19]

**Table 1.**
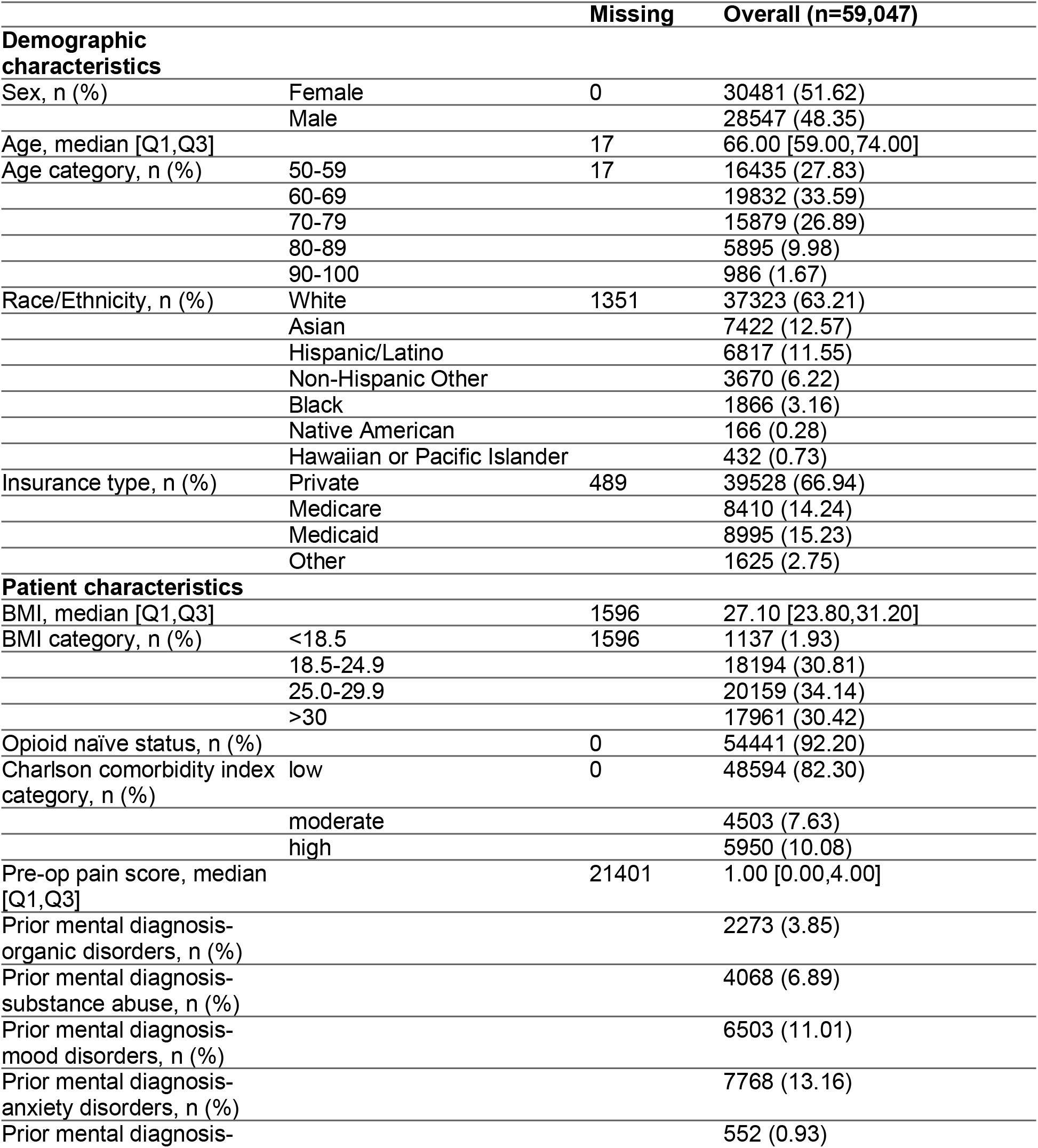
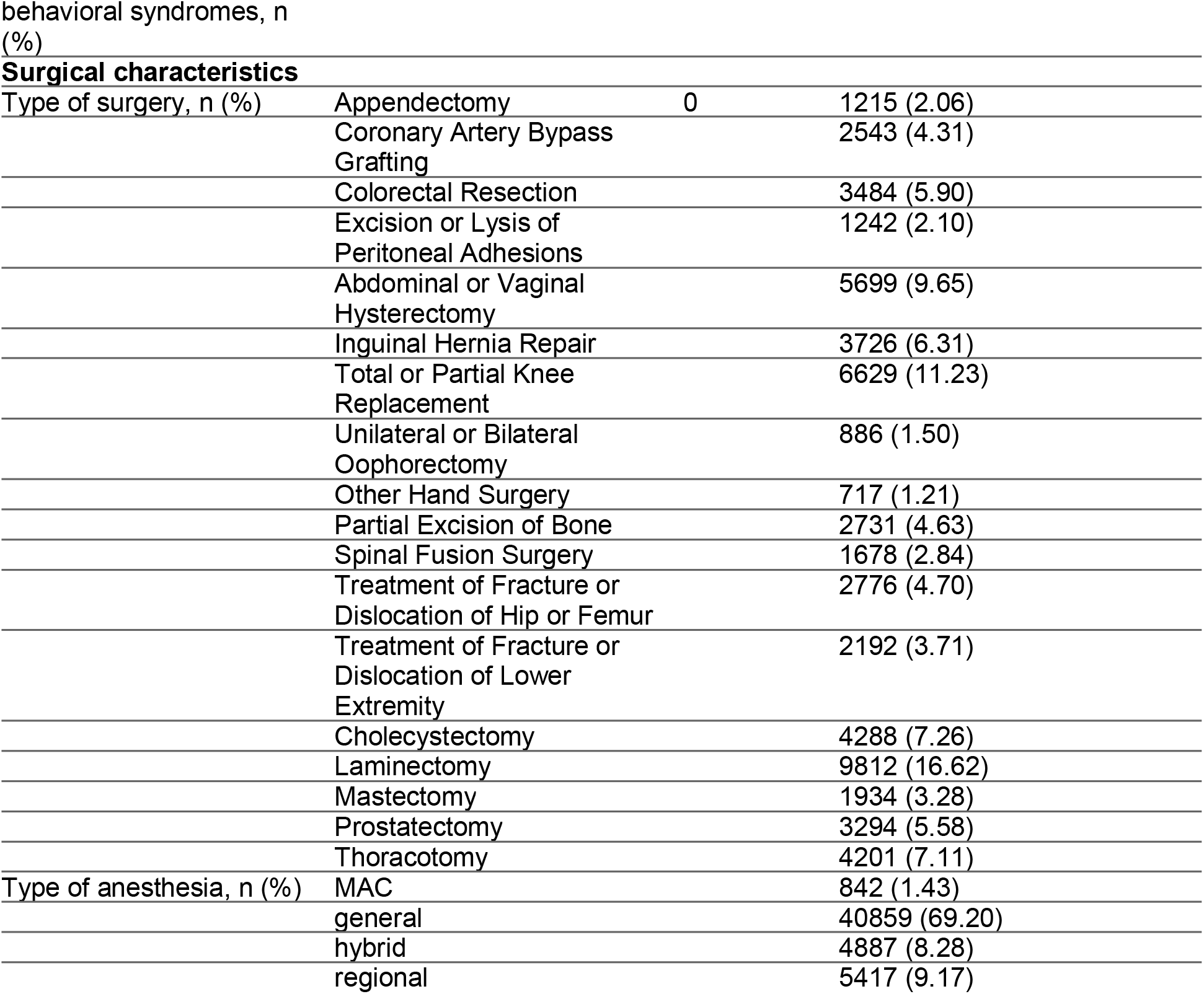
Baseline characteristics of Surgical Patients.

### 2.2 Model development

#### 2.2.1 Outcome

The primary outcome was post-surgical delirium, identified using ICD diagnostic codes (Table S1), documented positive Confusion Assessment Method (CAM) scores, and natural language processing of clinical notes (Method S1). We used NLP to supplement structured data because delirium is frequently underreported in coded fields. The secondary outcome was 30-day hospital readmission, defined as any inpatient admission within 30 days of discharge.

#### 2.2.2 Features

We categorized features into sensitive and clinical attributes. Sensitive attributes included sex, race/ethnicity, and insurance type (as a proxy for socioeconomic status). While sex can serve as either a sensitive or clinical attribute depending on context, we classified it as sensitive for this study. Clinical attributes included age, body mass index (BMI), pre-surgical pain score, Charlson Comorbidity Index (CCI), opioid-naïve status, mental health diagnoses, anesthesia type, and surgery type. We used the most recent BMI recorded prior to surgery. Pre-surgical pain scores were collected from a 180-day window before surgery through the day of surgery. The CCI was calculated using diagnoses from the two years preceding surgery. For descriptive analyses, we categorized CCI scores as low (0–2), moderate (3– 4), or high (≥5), while continuous scores were used in machine learning models. We defined opioid-naïve status as having no opioid prescription in the 6 months before surgery. Mental health diagnoses were identified using the five most frequent ICD-10 F-code categories in our cohort: mental organic disorders, substance use disorders, mood disorders, anxiety disorders, and behavioral symptoms (Table S1).

Anesthesia type was determined through rules-based natural language processing of operative and anesthesia procedure notes, with cases involving both general and regional anesthesia categorized as hybrid. Surgery type was extracted from surgical notes (Method S2).

All features were preprocessed using standard normalization and one-hot encoding. Missing values were imputed using median values.

#### 2.2.3 Models

We used an 80/20 train-test split. To address class imbalance, we applied the Synthetic Minority Oversampling Technique (SMOTE) to the training set. Models were developed using logistic regression and extreme gradient boosting (XGB). Model performance was assessed using accuracy, precision, recall, and area under the receiver operating characteristic curve (AUROC). All analyses were conducted using scikit-learn (v1.3.0) and XGBoost (v2.1.1) in Python. We also evaluated the contribution of each sensitive attribute to model performance and fairness by training models with and without individual sensitive attributes and examining the resulting disparities. We note that the models were not optimized for predictive performance or clinical deployment but serve as demonstration cases to illustrate the application of the EquiLense fairness auditing framework.

### 2.3 Fairness evaluation

#### 2.3.1 Fairness evaluation visualization framework

Group fairness was evaluated using Demographic Parity Difference (DPD), Equal Opportunity Difference (EOD), Equalized Odds (EO),[20] as well as the Error Distribution Disparity Index (EDDI), a novel group fairness metric designed for clinical settings with diverse group sizes and class imbalance.[18]

Individual fairness was assessed by measuring prediction consistency among clinically similar patients, with similarity defined using distance-based metrics. We calculated individual fairness metrics with both a narrow set of clinical variables (age, CCI, BMI) and the full feature set to examine how the definition of clinical similarity influences fairness estimates. For the full feature set, comorbidities included in the CCI were one-hot encoded. All metrics were computed on the test set.

Currently, there is currently no consensus on how to define clinical similarity in individual fairness assessments. Similarity may vary depending on the context, population, and prediction task. To support flexible exploration, our codebase allows users to specify their own similarity criteria, including variables, distance metrics, and thresholds. This enables customized fairness analyses across diverse clinical use cases.

#### 2.3.2 Mean Predicted Probability Difference (MPPD)

To evaluate fairness at both the group and individual levels, we developed a novel integrated metric, the Mean Predicted Probability Difference (MPPD). This metric quantifies inconsistencies in predicted probabilities between clinically similar patients who belong to different sensitive groups (Figure 1).

**Figure 1.**
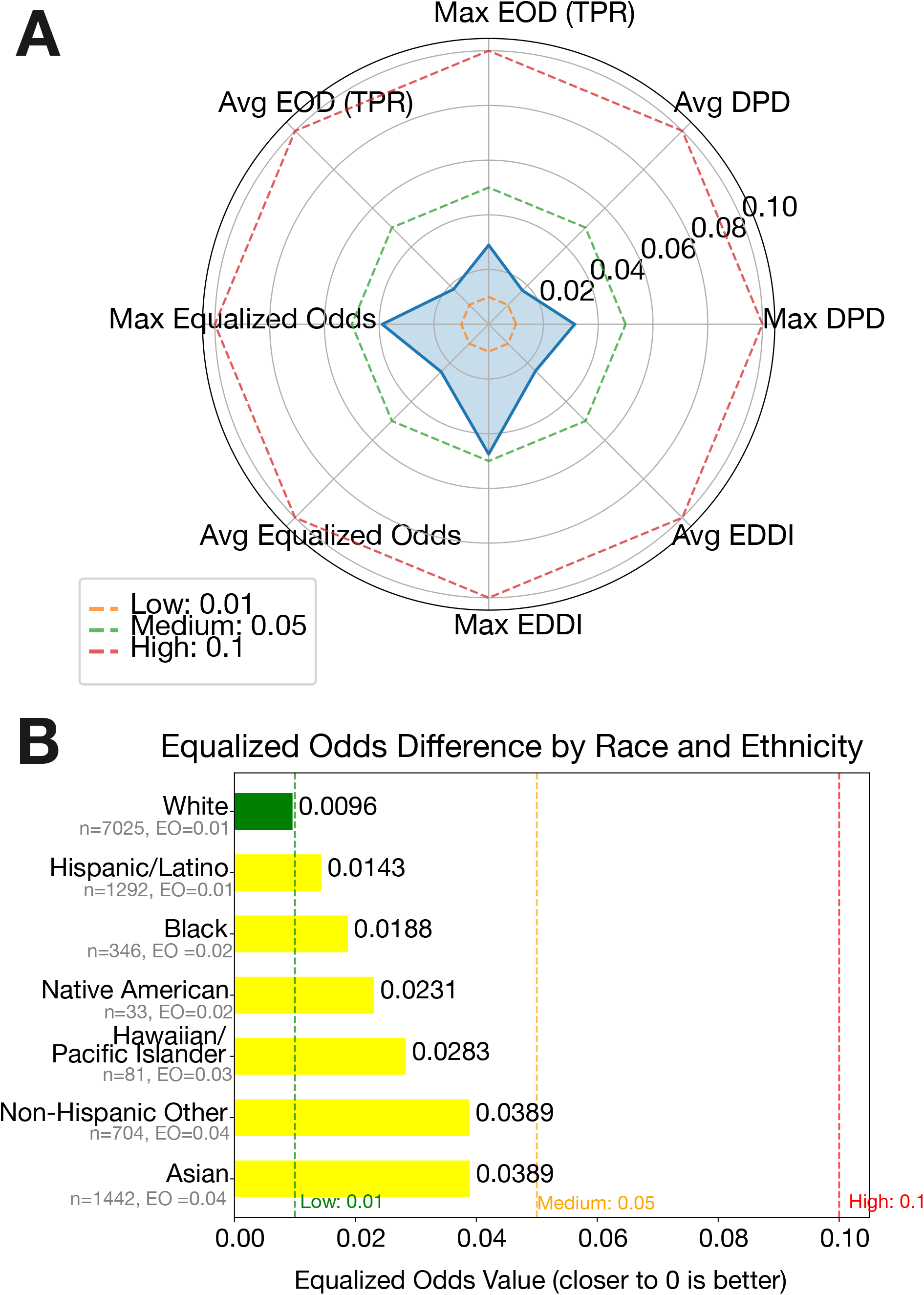
Conceptual overview of Mean Predicted Probability (MPPD) metric. Illustration of how MPPD is calculated. For each patient, predicted probabilities are compared with those of clinically similar patients from a different demographic group. In this example, the sensitive attribute is race. These differences are then averaged across group pairs to quantify disparities in prediction consistency.

We had two approaches: 1) reference-based and 2) pairwise. The reference-based approach begins by identifying the largest group within each sensitive attribute (e.g., Non-Hispanic White patients for race/ethnicity) as the reference group. For each patient in the reference group, we identify clinically similar individuals purely based on cosine similarity computed on clinical attributes. The similarity threshold, set to 0.01 by default but user-selectable, defines matched pairs. The set of clinical and social attributes used to define similarity can also be tailored based on user judgment. For example, for a Non-Hispanic White patient, they could have several matched pairs as long as the other patient’s clinical similarity distance is within 0.01 (the similar patients could be Non-Hispanic White patients, Non-Hispanic Asian patients, Hispanic patients, or from any of the race/ethnicity groups). They might also have no matched pair if there is no other patient within 0.01 of clinical similarity distance.

For each matched pair, we calculate the absolute difference in model predicted probabilities, and we aggregate these differences by group (for all of reference and non-reference groups) to obtain the average MPPD value for each group. This process captures whether patients from non-reference groups receive systematically different predictions despite having similar clinical profiles, thereby integrating elements of group fairness (via inter-group comparison) and individual fairness (via intra-clinical similarity).

To complement the reference-based approach and avoid assumptions about any one group being “fair” by default, we extended MPPD to a pairwise version. In this version, we performed bi-directional matching between all combinations of sensitive attribute groups, allowing identification of clinically similar patients across group-pairs (e.g., Black-White, Asian-Hispanic). For each group-pair, we computed the average absolute difference in predicted probabilities between matched patients, producing a matrix of pairwise MPPD scores. This symmetric comparison allows for a more comprehensive understanding of prediction consistency across all groups, including disparities between minority populations that would be overlooked in a reference-only framework.

Together, these two implementations of MPPD provide a flexible and interpretable method for uncovering fairness concerns in clinical prediction models. The interpretability of the framework comes from its use of clinically familiar features already involved in model development, while its flexibility lies in allowing users to define clinical similarity according to their own context (e.g., age, comorbidity burden, surgical type). This alignment with routine clinical reasoning makes the outputs easier for practitioners to understand and apply in practice.

### 2.4 External Benchmark Validation

To demonstrate MPPD against known ground truth, we applied it to two external datasets selected for their well-documented algorithmic bias: the COMPAS recidivism dataset (n=6172) and the UCI Adult Income dataset (n=32,561).[21, 22] For each dataset, an XGBoost classifier was trained on an 80/20 train-test split as a demosntration case. For COMPAS, the reference group was White defendants and similarity features included number of prior offenses, age bracket, and misdemeanor status. For UCI Adult Income, reference groups were White and Male. Similarity features included age, education level, capital gain, capital loss, hours worked per week, occupation, and work class.

## 3. RESULTS

### 3.1 Descriptive statistics

The cohort comprised 59,047 surgical patients who underwent 75,179 surgeries at a large academic medical center (Table 1 and Figure S1). Post-surgical delirium prevalence was 14.9% and 30-day readmission was 8.8%. The median age at surgery was 66 years, with 52% of patients being female. Most patients identified as Non-Hispanic White (63%), followed by Non-Hispanic Asian (13%) and Hispanic (12%). At surgery, 59% had private insurance (Table 1).

### 3.2 Model performance

For delirium prediction, the XGBoost model achieved an accuracy of 0.84, AUROC of 0.78, precision of 0.45, and recall of 0.26 (Table S2). Removing individual sensitive attributes (race/ethnicity, sex, or insurance) had minimal impact on model performance, with accuracy and AUROC remaining stable at 0.83 and 0.77, respectively. For 30-day readmission, XGBoost achieved an accuracy of 0.91, AUROC of 0.61, precision of 0.19, and recall of 0.02; excluding sensitive attributes again had negligible effects. Logistic regression performed worse across both outcomes. In both models, sensitive attributes had low-to-moderate feature importance and minimal effect on predictive performance when removed (Figure S2-S3).

### 3.3 Group and Indiviudal Fairness

Figure 2a presents a radar plot summarizing group fairness metrics for the delirium prediction model, using race as the sensitive attribute. Both maximum and average values are presented for DPD, EOD, EO, and EDDI, with disparity thresholds set at 0.01 (low), 0.05 (moderate), and 0.1 (high). EDDI shows the largest disparity, reflecting differences in prediction error distributions across groups. Figure 2b presents EO difference values by race and ethnicity. Asian and non-Hispaic other patients showed the highest EO differences (0.0389), in the moderate disparity range, while White patients had the lowest (0.0096).

**Figure 2.**
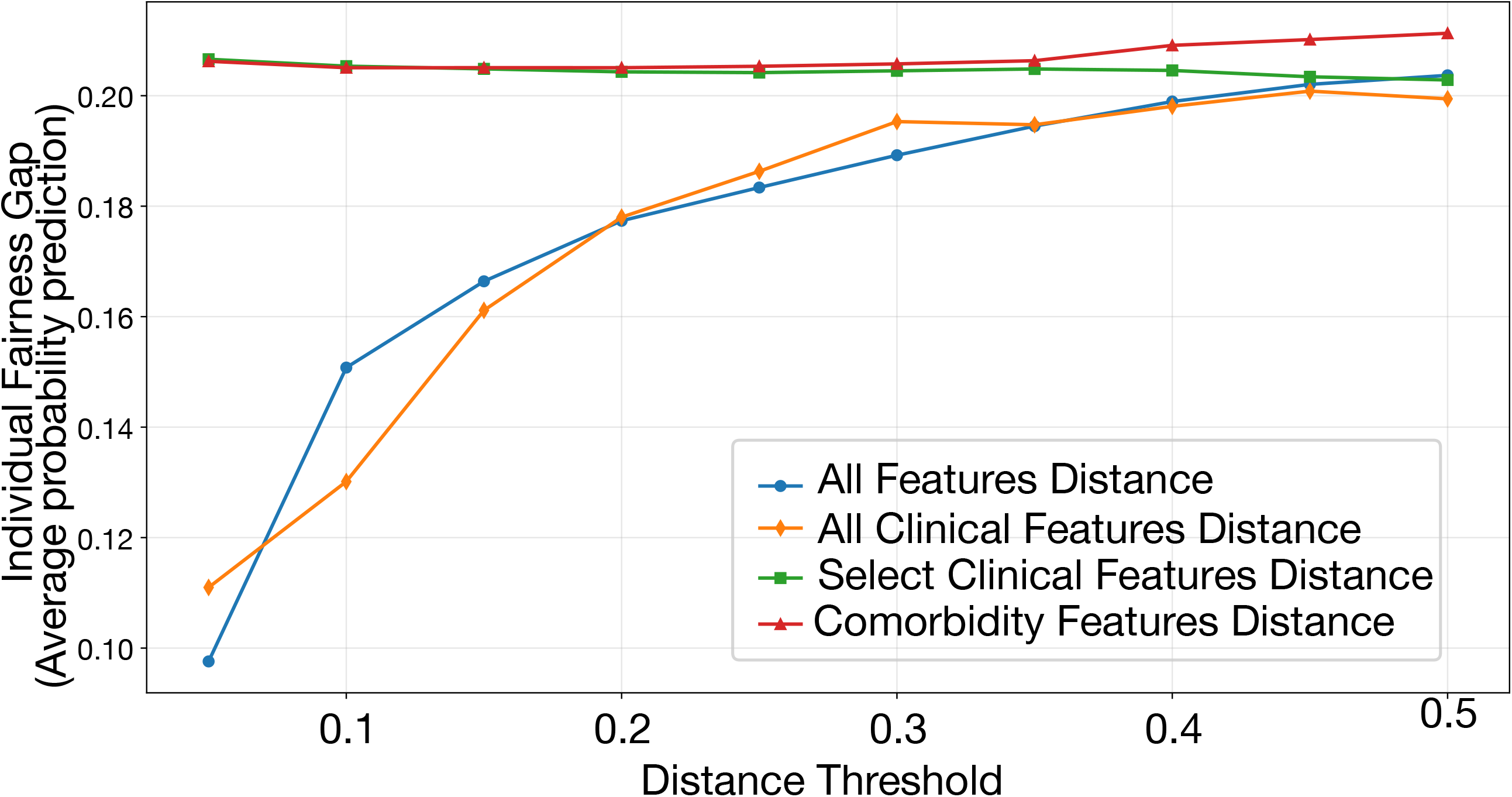
Visual summary of group fairness metrics in the delirium prediction model. Abbreviations: DPD, Demographic Parity Difference; EOD, Equal Opportunity Difference; EDDI, Error Distribution Disparity Index. Panel A displays a radar chart summarizing group fairness metrics across demographic groups, including DPD, EOD, Equalized Odds, and EDDI. Dashed lines indicate thresholds for low (0.01), medium (0.05), and high (0.1) disparity. Panel B presents Equalized Odds differences by race and ethnicity, highlighting subgroup disparities. Asian patients showed the highest disparity, and all groups fell within the low or moderate range. Test sample sizes and disparity values are shown for each group.

Figure 3 shows individual fairness gaps across feature sets and distance thresholds. Models using the full set of clinical features consistently achieved lower individual fairness gaps than narrower feature sets. These results illustrate how similarity definition influences fairness estimates, and presented as components of EquiLense’s integrated auditing toolkit.

**Figure 3.**
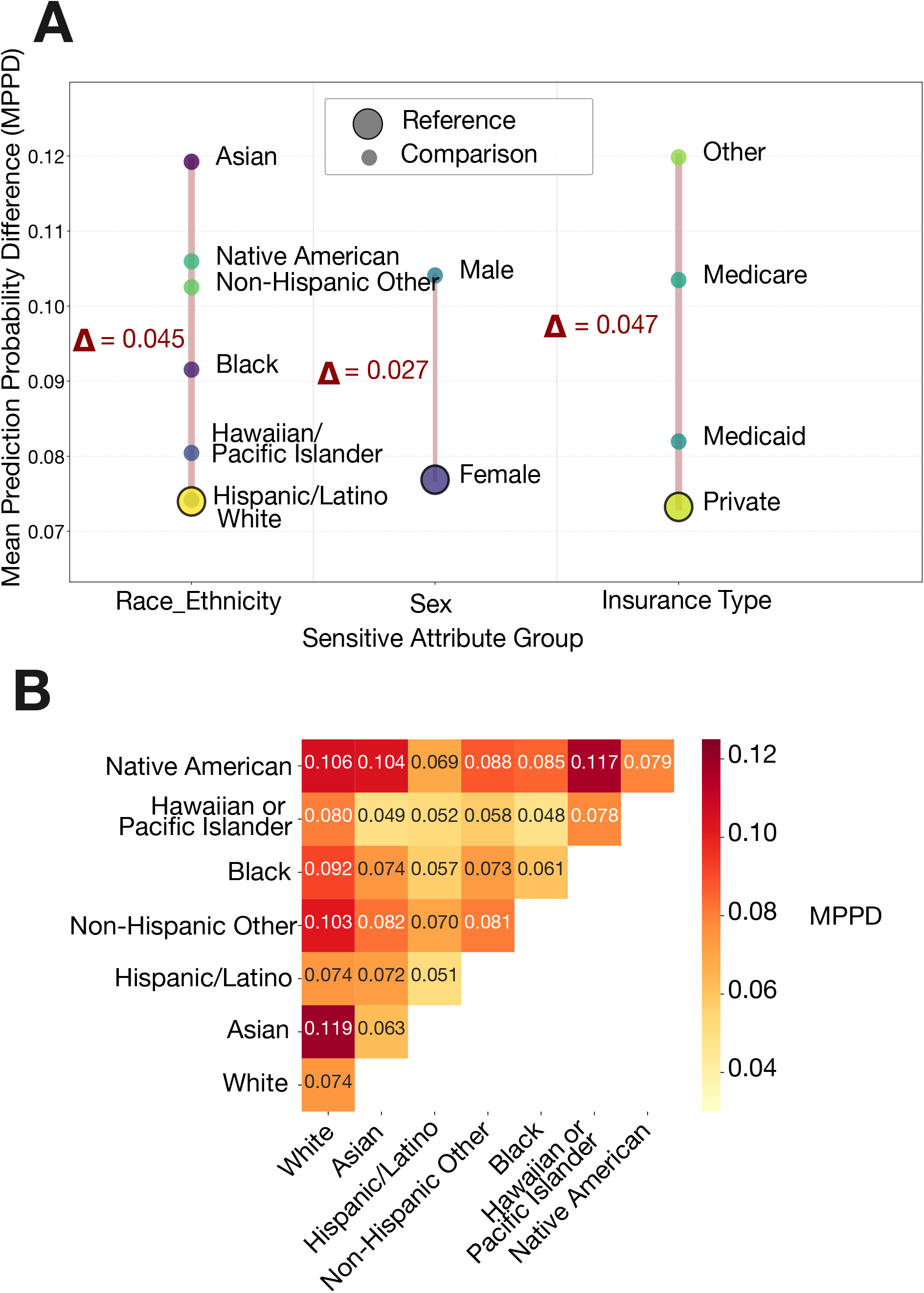
Individual fairness Scores by feature set and distance threshold. This displays individual fairness scores across varying clinical feature sets and distance thresholds. Individual fairness was assessed by calculating the average difference in predicted probabilities between clinically similar patients, with lower values indicating greater consistency (i.e., better fairness). Models using a comprehensive set of clinical features, including surgery and anesthesia type, showed lower fairness scores compared to those using narrower feature sets (such as age, BMI, Charlson Comorbidity Index, or comorbidity features alone).

### 3.4 Mean Predicted Probability Difference

To demonstrate how EquiLense can inform practical modeling decisions, we examined whether excluding sensitive attributes from model training affects MPPD scores.

Figure 4a presents MPPD values for the delirium model across sensitive attributes group. Among patients clinically similar to a White patient, Asian patients exhibited the largest prediction disparity (MPPD White-Asian = 0.119, MPPD White-White = 0.074, □ = 0.045), indicating lower individual fairness for that group. Excluding race and ethnicity from model training reduced these disparities across most group pairs (Figure S4), suggesting that feature inclusion directly influences prediction consistency between demographically different but clinically similar patients.

**Figure 4.**
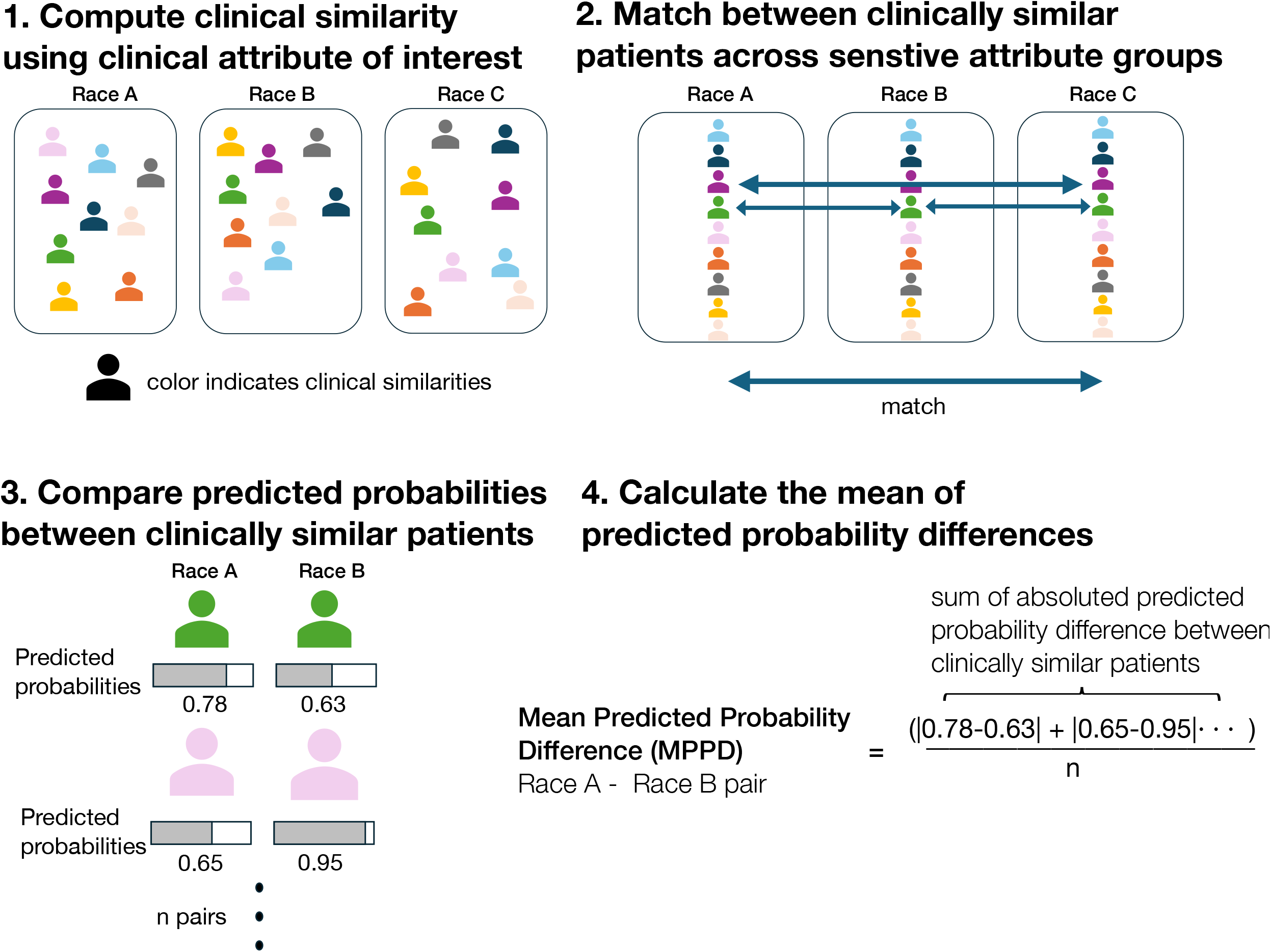
MPPD across sensitive attribute groups. Panel A shows MPPD values comparing predicted probabilities among clinically similar patients across sensitive attribute groups, including race/ethnicity, sex, and insurance type. The largest disparity was observed between Asian and White patients (White-Asian: 0.119 vs. White-White: 0.074; Δ = 0.045), highlighting prediction inconsistencies. Panel B displays the full pairwise MPPD matrix across racial and ethnic groups, with each cell reflecting the MPPD between matched patients from the corresponding group pair. Within-group MPPD values are non-zero because each patient is compared to another clinically similar patient from the same group.

Figure 4b presents the full pairwise MPPD matrix. Within-group MPPD values (e.g. White-White) are non-zero because each patient is compared to another clinically similar patient from the same group. When race and ethnicity were excluded from model features (Figure S5), MPPD scores decreased substantially across most group pairs. The exception was the Black-Native American pair; this estimate should be interpreted with caution given the small Native American test sample size (n=32), a recognized limitation in health disparities research. Similar trends were observed in the 30-day readmission model.

### 3.5 External Benchmark Validation

MPPD detected known bias in both external datasets. In COMPAS, African American defendants received the highest MPPD score (0.1122), consistent with the well-documented pattern of disproportionately elevated recidivism risk scores assigned to Black defendants relative to White defendants with similar criminal histories.[23, 24] In the UCI Adult Income dataset, female workers received the highest MPPD score (0.2363), consistent with documented gender bias in income prediction.[25] Across both datasets, MPPD assigned the highest disparity scores to groups with known algorithmic disadvantage, supporting its validity as a fairness screening metric.

## 4. DISCUSSION

We present EquiLense, a model-agnostic, post-hoc auditing tool that addresses a gap in current fairness evaluation: the lack of a practical, accessible tool that integrates both group-level and individual-level fairness assessment for already-deployed clinical prediction models. While existing tools address one perspective or require model retraining, EquiLense integrates group fairness evaluation, individual fairness assessment, and MPPD, its novel methodological contribution, into a single framework that requires no model retraining and applies to any clinical prediction model. Applied to surgical outcome models, we demonstrate that strong overall predictive performance does not preclude systematic prediction inconsistencies across demographic groups. External benchmark demonstration on COMPAS and UCI Adult Income confirms that MPPD detects meaningful disparities when bias is known to exist.

Current fairness evaluation approaches remain fragmented. Although fairness and bias mitigation strategies in clinical AI are increasingly studied, consensus on what constitutes fairness remains elusive, with definitions that often conflict and require trade-offs. [5] Different fairness metrics serve different purposes, and no single metric is sufficient on its own. Group fairness metrics identify systemic disparities but are insensitive to individual-level variation within groups, while individual fairness metrics have seen limited adoption in healthcare because similarity definitions are inherently context-dependent, varying across clinical tasks, populations, and outcomes.[11] Evaluating fairness comprehensively requires applying multiple metrics that reflect different fairness perspectives, as different metrics can lead to conflicting conclusions and the choice of metric should align with the clinical context and ethical goals of the specific use case.[5] EquiLense addresses a key dimension of this fragmentation by integrating group fairness evaluation, individual fairness assessment, and MPPD into a single tool, enabling fairness assessment that captures both population-level patterns and patient-level inconsistencies. The tool allows users to specify clinically grounded similarity criteria, aligning the evaluation with clinical reasoning relevant to the prediction task at hand. MPPD values represent prediction differences between clinically similar patients, allowing users to assess both whether disparities exist and their magnitude, supporting practical decision-making about model deployment and feature selection.

MPPD is designed to complement, not replace, existing fairness approaches. It compares real, observed patients who are already clinically similar, grounding the fairness assessment in observed data. Compared to CF-based approaches, MPPD is computationally straightforward and does not require causal group specification, synthetic data generation, or model training, lowering the barrier for practical fairness auditing in clinical settings. Its assumptions concern which clinical variables define meaningful similarity. These are explicit, clinically interpretable, and directly adjustable by practitioners. Studies suggest that fairness should be addressed throughout all phases of model development and deployment, not only at a single stage.[5] MPPD is particularly well-suited for post-hoc auditing of already-deployed models, a stage that training-time approaches cannot address. In this way, EquiLense and FairEHR-CLP,[18] our CF-inspired in-processing method, address complementary stages of the clinical AI lifecycle. FairEHR-CLP evaluates and addresses fairness during model development, while EquiLense detects and surfaces disparities after deployment through MPPD, its individual-to-group fairness bridging metric.

MPPD is intended as a screening and auditing metric. A high MPPD between two demographic groups indicates that clinically similar patients from those groups receive systematically different predictions. Whether this reflects algorithmic bias, an imperfect similarity specification, or clinically appropriate distinctions requires further investigation and clinical judgment. This interpretive challenge is shared with group fairness metrics, which similarly flag disparities without resolving their cause. EquiLense supports this investigative process by enabling sensitivity analyses across similarity thresholds, feature sets, and modeling choices.

To illustrate how EquiLense can inform feature selection decisions, we examined the effect of excluding sensitive attributes from model training on MPPD scores. We observed that excluding sensitive attributes, such as race/ethnicity and insurance type, from model training resulted in lower MPPD scores, indicating improved fairness in prediction consistency across demographic groups. In our demonstration, removing these attributes reduced disparities between clinically similar patients without affecting overall model performance, though MPPD estimates for the Black-Native American pair should be interpreted with caution given the small Native American test sample size (n=32), a recognized limitation in health disparities research. These sensitive attributes also had low feature importance, suggesting minimal contribution to predictive accuracy. However, algorithmic bias can arise from multiple sources, including data imbalance, labeling variation, and broader structural inequities.[4, 11] There is rarely a single solution. Removing a sensitive attribute is one possible approach but does not guarantee improved fairness in all cases.[5] EquiLense enables users to evaluate these trade-offs by testing how different modeling choices affect both fairness and performance.

A key strength of EquiLense is its adaptability to diverse clinical contexts. Fairness goals and clinical priorities vary across healthcare settings, so the framework allows users to tailor evaluations accordingly. For example, age may be treated as a clinical predictor in surgical risk stratification but as a sensitive attribute in fairness assessments, while sex may serve as a biological covariate or as a sensitive attribute depending on the use case. Users can specify which variables define clinical similarity, which are treated as sensitive attributes, and adjust similarity thresholds and metric selection. This flexibility ensures fairness evaluation aligns with the specific priorities of each clinical application.

Our approach is conceptually related to counterfactual fairness methods but differs in how it operationalizes the fairness principle. CF evaluates whether a model’s prediction would change for a hypothetical version of the same individual with a different demographic identity. This requires specification of a causal graph that captures how changes in sensitive attributes propagate through other features.[13, 26, 27] CF-inspired approaches, including our prior work FairEHR-CLP,[18] may approximate this through synthetic counterpart generation without full causal graph specification, but still require assumptions about which features would change under a different demographic identity.[14] Such assumptions are rarely verifiable in complex clinical settings due to unmeasured confounders, treatment heterogeneity, and documentation biases in EHR data. [14, 15] While MPPD also relies on assumptions about which clinical variables appropriately define similarity, these are grounded in observable clinical features and directly adjustable by practitioners based on domain knowledge. We acknowledge that CF-based approaches may account for certain confounding pathways that MPPD does not by modeling causal structure. However, such causal assumptions are rarely verifiable in EHR settings, and concerns about structural bias in EHR-derived features apply equally to all EHR-based research. In contrast to CF, EquiLense matches real, observed patients from different demographic groups who are clinically similar, rather than hypothetical or synthetic counterparts. This grounds the fairness assessment in observed data, making its assumptions explicit and inspectable. A further strength of the pairwise comparison approach is that it enables symmetric evaluation across all group combinations without relying on a fixed reference group, surfacing disparities between minority populations that reference-based approaches would overlook.

Despite its advantages, our approach has several limitations. First, MPPD is not a causal measure and cannot determine whether observed differences in predicted probabilities reflect unfair bias or clinically appropriate distinctions. It provides a systematic way to surface inconsistencies that warrant further investigation but requires clinical judgment to interpret. Second, fairness evaluation accuracy depends on how well clinical similarity is defined. Incomplete or noisy feature data may lead to mismatches that distort estimates. Defining similarity inherently involves subjective decisions about which features to include and how to weigh them, and fairness estimates can be sensitive to similarity thresholds. Additionally, chosen similarity variables such as CCI and BMI may themselves reflect structural inequities in healthcare access and documentation, a limitation shared by all EHR-based research. While EquiLense allows flexible specification of these parameters, users must carefully consider what constitutes meaningful similarity for their specific task. Future work should investigate how threshold selection affects fairness estimates and whether standard guidelines can be developed. Third, we did not explore interactions between demographic and clinical predictors, which may reveal additional sources of bias. Fourth, while EquiLense identifies disparities in model outputs, it does not assess downstream clinical consequences such as differences in care decisions or health outcomes. Finally, as a diagnostic tool, EquiLense does not include mitigation strategies.

Future work should focus on validation of EquiLense on across a broader range of clinical prediction tasks and patient populations, development of integrated approaches combining fairness auditing with bias mitigation, and establishment of clinical thresholds for meaningful MPPD values across different prediction tasks. Further investigation into how similarity threshold selection affects fairness estimates, and whether standardized guidelines can be developed across clinical contexts, represents an important direction. Formal usability testing with clinical stakeholders would also be a valuable next step to evaluate whether EquiLense’s design choices effectively support practical fairness auditing in real-world healthcare settings.

## 5. CONCLUSION

We introduced EquiLense, a flexible and model-agnostic tool for auditing fairness in clinical prediction models. By integrating group fairness evaluation, individual fairness assessment, and MPPD into a single post-hoc framework, EquiLense addresses a key gap in current fairness evaluation: the lack of a practical tool that captures both population-level disparities and patient-level prediction inconsistencies without requiring model retraining. Using post-surgical delirium and readmission models, we demonstrated how EquiLense surfaces fairness concerns, supports sensitivity analyses, and guides decisions about model design and feature inclusion. External validation on COMPAS and UCI Adult Income confirms that MPPD detects meaningful disparities when bias is known to exist. As clinical AI systems become more deeply integrated into care delivery, comprehensive and interpretable fairness auditing frameworks will be essential for ensuring that these systems advance rather than undermine health equity.

## Acknowledgments

None

## Author contributions

JX: Conceptualization, Resources, Data Curation, Software, Formal Analysis, Investigation, Visualization, Data Interpretation, Methodology, Writing - original draft, Writing - review and editing; Y-MH: Conceptualization, Resources, Data Curation, Software, Formal Analysis, Investigation, Visualization, Data Interpretation, Methodology, Writing - original draft, Writing - review and editing; SK: Formal Analysis, Investigation, Writing – original draft; ID: Methodology, Data Curation, Software, Writing - review and editing; SLJ: Investigation, Writing - review and editing; MP: Methodology, Data Interpretation, Writing - review and editing; CC: Data Curation, Data Interpretation, Methodology, Writing - review and editing; TH-B: Conceptualization, Supervision, Writing - review and editing, Administrative, technical, or material support; JX, Y-MH, ID, MP and TH-B had full access to the primary cohort data in the study and verified the data. All authors read and approved the final manuscript.

## Data availability

The data underlying this article cannot be shared publicly due to patient privacy protections and institutional data use agreements. Deidentified data may be available upon reasonable request to the corresponding author and with appropriate data use agreements and institutional review board approvals. Code implementing the EquiLense framework is publicly available at https://github.com/su-boussard-lab/equilense_fairness

## Alt text

Figure 1:Alt text: Diagram illustrating how MPPD is calculated by comparing predicted probabilities between clinically similar patients from different racial groups, with differences averaged across group pairs to quantify prediction disparities.

Figure 2:Alt text: Panel A shows a radar chart summarizing group fairness metrics including DPD, EOD, EO, and EDDI for the delirium prediction model with disparity thresholds marked. Panel B shows a bar chart of Equalized Odds differences by race and ethnicity with Asian patients showing the highest disparity.

Figure 3:Alt text: Line graph showing individual fairness scores across different clinical feature sets and distance thresholds, with models using comprehensive feature sets showing lower fairness gaps than narrower feature sets.

Figure 4:Alt text: Panel A shows bar charts of MPPD values across race/ethnicity, sex, and insurance type sensitive attribute groups. Panel B shows a heatmap of pairwise MPPD scores across all racial and ethnic group combinations.

