## Supplementary Materials for "Integrating Group and Individual Fairness Auditing in Clinical AI: A Post-Hoc, Model-Agnostic Approach"

### **Table of Contents**

**Supplementary Tables**

Table S1 ICD diagnostic codes for post-surgical delirium phenotype identification

Table S2 Descriptive statistics of post-surgical patients

Table S3 Postoperative delirium and 30 day readmission prediction model performance

Table S4 Regex string patterns used in Method S1

**Supplementary Figures**

Figure S1 Cohort selection figure adapted from prior work

Figure S2 Feature importance of XGBoost Model for postoperative delirium prediction

Figure S3 Feature importance of XGBoost Model for postoperative 30 day readmission prediction model

Figure S4 Reference-based MPPD after Excluding Sensitive Attributes from Model Training

Figure S5 Pairwise MPPD after Excluding Sensitive Attributes from Model Training

**Supplementary Methods**
Method S1 Delirium phenotyping via NLP model development

Method S2 List of surgical types

### **Supplementary Tables**

Table S1 ICD diagnostic codes for post-surgical delirium phenotype identification

| **ICD9** | 293, 293.1, 292.81, 290.11, 290.3, 290.41, 293.9, 780.09, 293.81, 293.82, 293.83, 293.89, 290.12, 290.13, 290.43, 292.11, 292.12, 292.2, 780.02, 290.2, 290.42, 290.8, 290.9, 292, 292.82, 348.31, 348.39, 349.82, 780.97 |
| --- | --- |
| **ICD10** | F05, F11.121, F11.221, F13.121, F13.221, F13.231, F11.921, F13.921, R41.0. F19.921, F03.90, F01.51, F06.8, R40.0, F06.2, F06.0, F06.30, F03.90, F19.950, F19.951, R40.4, F19.97, G93.49, G92, R41.82. |

Table S2 Postoperative delirium and 30 day readmission prediction model performance

| **Task** | **Model** | **Accuracy** | **Precision** | **Recall** | **AUROC** |
| --- | --- | --- | --- | --- | --- |
| **Delirium** | Original | 0.84 | 0.45 | 0.29 | 0.78 |
| **Delirium** | No race and ethnicity | 0.83 | 0.42 | 0.28 | 0.77 |
| **Delirium** | No sex | 0.84 | 0.43 | 0.27 | 0.77 |
| **Delirium** | No insurance | 0.83 | 0.40 | 0.02 | 0.77 |
| **Readmission** | Original | 0.91 | 0.17 | 0.03 | 0.60 |
| **Readmission** | No race and ethnicity | 0.91 | 0.19 | 0.02 | 0.60 |
| **Readmission** | No sex | 0.91 | 0.17 | 0.02 | 0.59 |
| **Readmission** | No insurance | 0.91 | 0.21 | 0.29 | 0.58 |

Table S3 Regex string patterns used in Method S1

| **Component** | **Terms/Patterns** |
| --- | --- |
| **Inclusion**  **terms** | ['CAM Score Yes', 'CAM Score Positive','delir(ium\|ious\|ation)?', 'delirium state','(acute (onset\|change in mental status)\|mental status change\|fluctuating mental status\|(altered\|waxing and waning) mental status)\ (inatten(tive\|tion)\|distract(ed\|ion)\|confus(ed\|ion)\|disorient.*)\ (disorganized thinking\|altered level of consciousness\|letharg(y\|ic)\|fluctuating  arousal\|agitat(ed\|ion)\|encephalopathy)'] |
| **Exclusion**  **terms** | ['Dementia','alzheimer', 'schizophrenia', 'bipolar disorder', 'CAM score no', 'CAM score negative'] |
| **Negation**  **cues** | ['no ','not ', "n't", 'negative', 'never', 'denies', 'deny', 'risk','assessment', 'prevention'] |

### **Supplementary Figures**

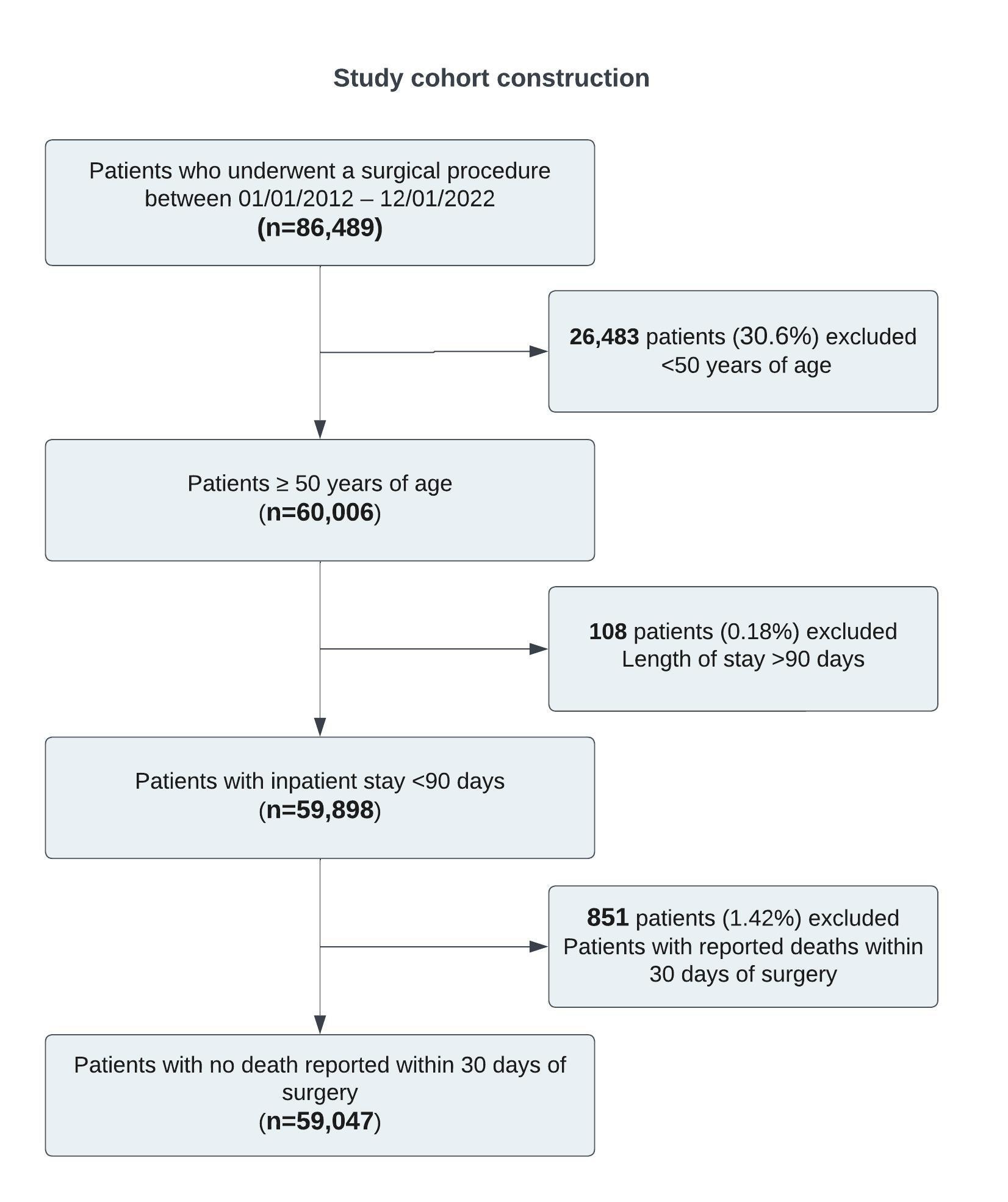

Figure S1 Cohort selection figure adapted from prior work (citation will be provided for camera-ready version)

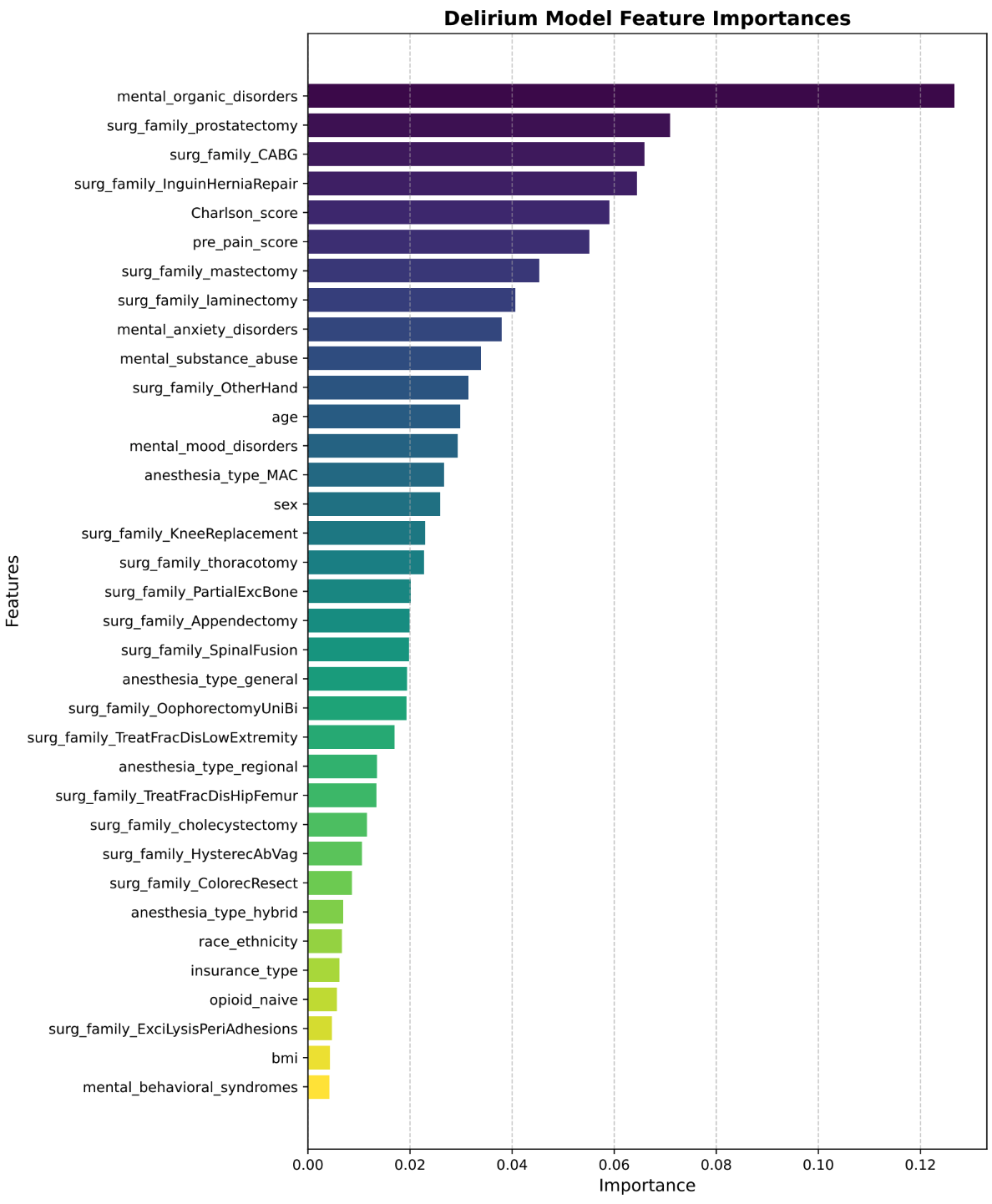

Figure S2 Feature importance of XGBoost Model for postoperative delirium prediction

This figure shows the relative importance of features used in the delirium prediction model. Clinical predictors such as Charlson Comorbidity Index (CCI), mental and behavioral disorders, pre-operative pain scores, and specific surgical procedures (e.g., CABG, hernia repair, prostatectomy) were among the most important features. Sensitive attributes such as race/ethnicity and insurance type had low importance in this model.

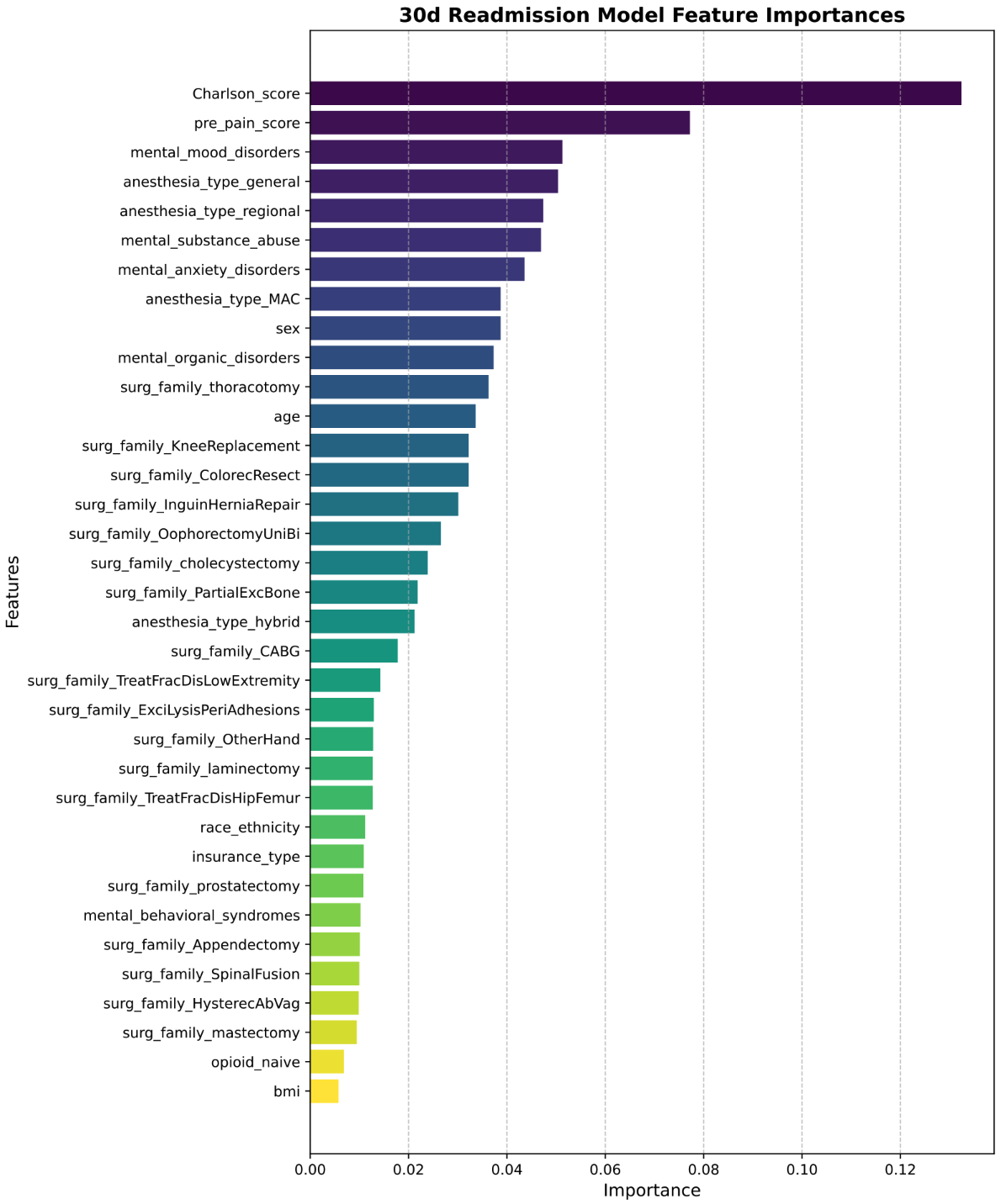

Figure S3 Feature importance of XGBoost Model for postoperative 30 day readmission prediction model

This figure displays the relative importance of features used in the readmission classification model. Key predictors included Charlson Comorbidity Index (CCI), anesthesia type and pre-existing mental health conditions. Sex showed moderate importance, while other sensitive features such as race/ethnicity and insurance type had low influence on model predictions.

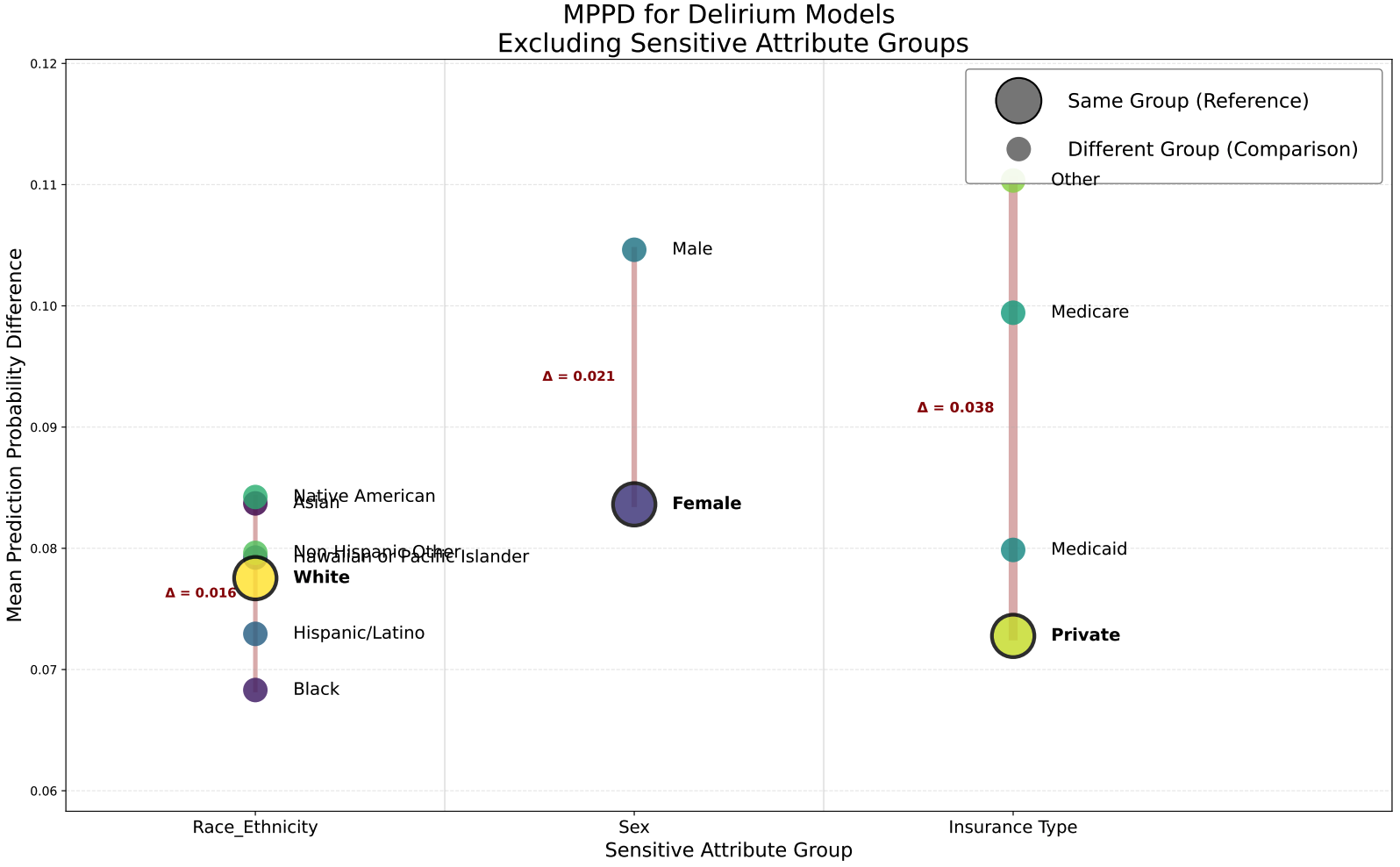

Figure S4 Reference-based MPPD after Excluding Sensitive Attributes from Model Training

Reference-based mean predicted probability differences (MPPD) across sensitive attributes

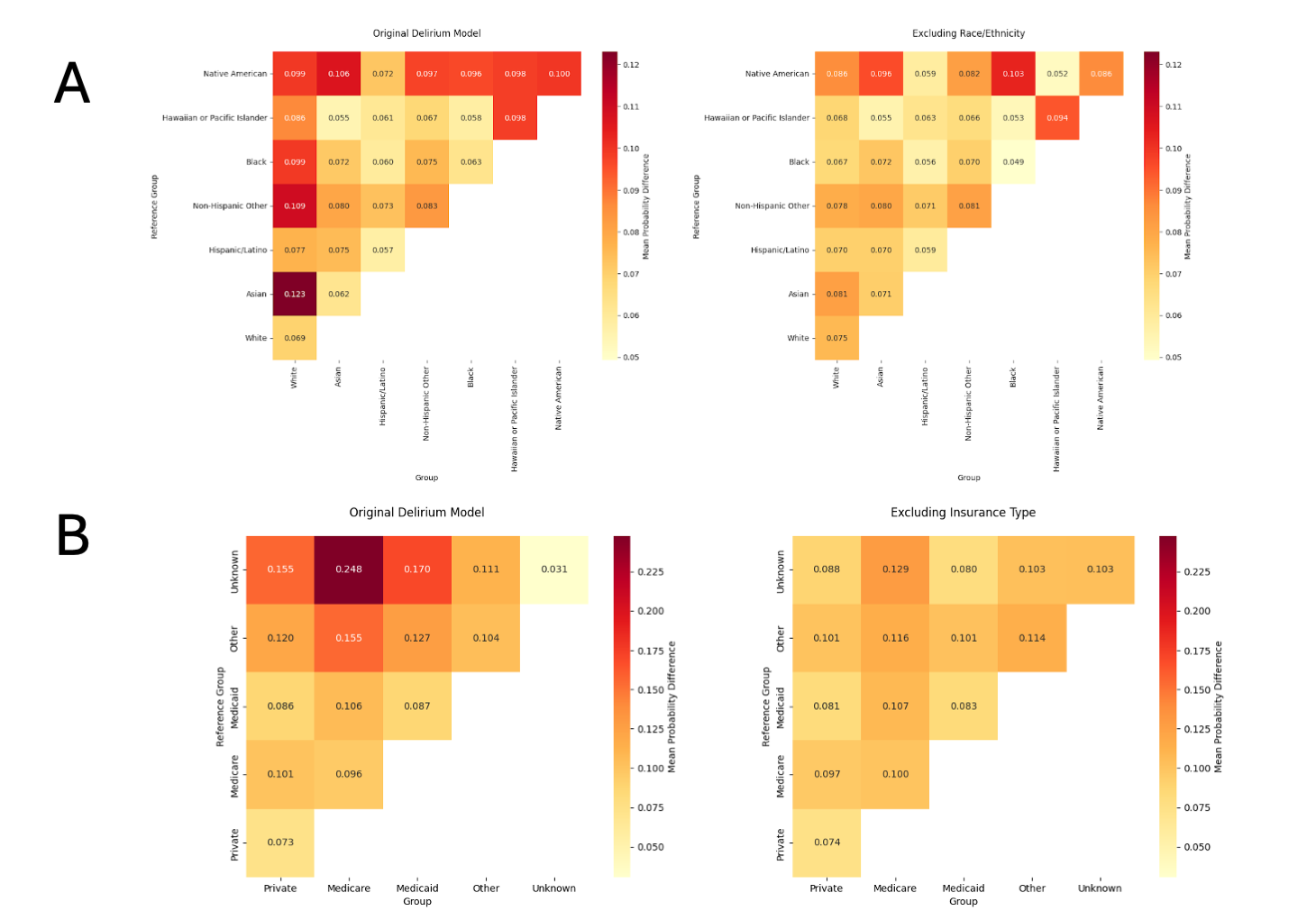

Figure S5 Pairwise MPPD after Excluding Sensitive Attributes from Model Training

(A) Pairwise mean predicted probability differences (MPPD) across racial and ethnic groups in the original model (left) and after excluding race/ethnicity from the training features (right). (B) Pairwise MPPD across insurance groups in the original model (left) and after excluding insurance type from the training features (right). Each cell represents the average difference in predicted probabilities between clinically similar patients from different groups. Excluding sensitive attributes reduced most group-level disparities in predicted probabilities.

### **Supplementary Methods**

Method S1 Delirium phenotyping via NLP model development

To train and evaluate our NLP model for delirium detection, we first constructed a labeled dataset using structured and unstructured EHR data. Patients were included if they met at least one of three criteria: presence of delirium-related ICD codes, positive CAM assessments in flowsheets, or keyword matches based on a regular expression (regex) search in clinical notes. Patients were labeled as positive if they were regex-positive and either ICD- or flowsheet-positive. From these patients, notes containing regex matches were extracted and filtered to include only those documented within a three-day window following the initial regex match. These were labeled as positive notes. Negative notes were sampled from patients who were negative for all three indicators to achieve a 40/60 positive-to-negative balance for model training and validation. For testing, a separate set of regex- and code-positive patients was sampled, along with a set of patients negative for all criteria, targeting a 20/80 positive-to-negative balance. Clinical experts manually annotated a subset of test notes into four categories: postoperative delirium, delirium risk, no delirium, and other (e.g., documentation of management or precautions). These labels were used to evaluate model performance. Multiple BERT-based models were trained using combinations of positive notes, hard negatives (e.g., notes with negated mentions), and easy negatives (notes with no relevant terms). The final model was applied to unstructured notes in the full cohort to identify additional delirium-positive documentation not captured by structured data or keyword searches alone.

Method S2 List of surgical types

Appendectomy, CABG, Colorectal Resection, Excision/Lysis Peritubal Adhesions, Hysterectomy (Vaginal/Abdominal), Inguinal Hernia Repair, Knee Replacement, Oophorectomy, Partial Excision of Bone, Spinal Fusion, Fracture Hip or Femur, Fracture Lower Extremity, Colecystectomy, Laminectomy, Mastectomy, Prostatectomy, Thoracotomy
